# SARS-CoV-2 seroprevalence in Chattogram, Bangladesh before the Delta surge, March-June 2021

**DOI:** 10.1101/2021.07.16.21260611

**Authors:** Taufiqur Rahman Bhuiyan, Juan Dent Hulse, Sonia T. Hegde, Marjahan Akhtar, Md. Taufiqul Islam, Zahid Hasan Khan, Ishtiakul Islam Khan, Shakeel Ahmed, Md Mamunur Rashid, Rumana Rashid, Emily S. Gurley, Tahmina Shirin, Ashraful Islam Khan, Andrew S. Azman, Firdausi Qadri

## Abstract

In a representative serosurvey conducted March–June 2021, 64.1% (95%CrI 60.0– 68.1%) of Sitakunda subdistrict (Bangladesh) had anti-SARS-CoV-2 IgG antibodies after adjusting for age, sex, household clustering and test performance. Before the surge of Delta, most of the population had been infected despite low incidence of virologically-confirmed COVID-19.

## Text

Since the start of the COVID-19 pandemic, Bangladesh has reported more than 1.55 million cases and 27,270 deaths (*1*). Reported mortality and incidence rates appear to be substantially lower than many other countries. Without population-based seroprevalence estimates, it is difficult to know whether these differences are due to limited surveillance and healthcare seeking or true differences in incidence resulting from interventions or differing biological responses to infection. In early March 2021, cases across Bangladesh began to rise at the same time the Delta variant was detected in neighboring India. Publicly available sequencing data (*2*) indicates the SARS-CoV-2 Delta variant was first detected in the Chattogram region of Bangladesh in mid-May 2021 and since July 1, 2021, 99% (n=98/99) of the submitted viral genomes have been the Delta variant, similar to national trends.

## The Study

We conducted a representative serosurvey to understand the prevalence of anti-SARS-CoV-2 IgG antibodies in the Sitakunda subdistrict (Chattogram district) of Bangladesh, which has a significant urban-rural gradient starting from Bangladesh’s second largest city, Chattogram. The survey was conducted in two periods, from 27-March to 13-April and from 23-May to 13-June (after Eid al-Fitr), due to a national COVID-19-related lockdown. We used two-stage sampling based on digitized satellite imagery, where we first selected a 1km^2^ grid-cell proportional to the estimated number of households in each grid, then randomly selected a structure weighted by whether it was multi- or single-story. We attempted to enroll all individuals ≤1 years old in each household.

We tested participant serum for total antibodies (IgG, IgM and IgA) against the S1-RBD of SARS-CoV-2 using the Wantai SARS-CoV-2 Ab ELISA kit (Beijing Wantai Biological Pharmacy Enterprise Co., China) following manufacturer instructions. We corrected seroprevalence estimates for imperfect test performance, household clustering and individual-level covariates (e.g., age) using a previously documented Bayesian modeling approach and post-stratified results to match the target population structure (*3*). This study was approved by the icddr,b Research and Ethics Review Committees and the Johns Hopkins Bloomberg School of Public Health Institutional Review Board. Additional methodological details, including details on sampling and enrollment, are in the supplement, and code and data to reproduce analyses are at https://github.com/HopkinsIDD/sitakunda-sarscov2-round1.

Given the limited data on the immunoassay’s performance in South Asia and more generally on its performance months after infection, we conducted a validation study to estimate test performance. We estimated the sensitivity and the specificity of the assay by testing samples from 214 healthy participants from a 2014 cholera vaccine study (pre-pandemic) and 81 samples from 52 symptomatic PCR-confirmed SARS-CoV-2 infections. None of the positive controls were hospitalized for COVID-19 nor vaccinated, and samples were collected 3 to 275 days post-symptom onset. We estimated a specificity of 99.1% (95%CI 96.7-99.9%, n=212/214) and sensitivity of 87.7% (95%CI 78.5-93.9, n=71/81) for detecting previous infection with little evidence of decreasing sensitivity with time since infection (**Supplementary Table 4**).

We enrolled 580 households and 2,307 individuals who provided a blood sample. A majority of participants were female (54%, n=1235/2307), with a median age of 28 (IQR: 16–45) years. Most participants reported working at home (37%), going to school (29%), or conducting business outside of their home (20%) as their main occupation in the month prior to enrollment. Among all study participants, 22 individuals (0.95%) reported ever having been tested for COVID-19 including 3 with positive results (3/3 were seropositive). One hundred and twenty-five (5.4%) reported being vaccinated with at least one dose of the Covishield (ChAdOx1, n=117) or Pfizer (n=1) SARS-CoV-2 vaccines. As of 19-June-2021, 6 days after the end of the survey, 6.0% of the entire Chattogram district population is reported to have received at least one dose of any vaccine including 4.6% with two doses (*4*).

There were 1,443 (63%) seropositive individuals. Nearly all (98%) who reported having been partially (n=47/49) or completely vaccinated (n=75/76) were seropositive. At least one individual was seropositive in 85% of enrolled households and, on average, 62% of participants in each household were seropositive. We estimate that 31% of the total variability in seropositivity in the community is attributed to variation in seropositivity between households (intraclass correlation coefficient: 0.31; 95%CI: 0.27–0.36). We found evidence of a gradient in seropositivity by population density, with those living in areas of higher population density being significantly more likely to be seropositive; 69% of individuals living in the most population dense areas were seropositive compared with 52% of individuals living in the least population dense areas (p<0.0001, Supplemental Table 1). We found similar results using alternative metrics related to urbanicity (Supplemental Table 1).

Adjusting for age, sex, household clustering, and test performance we estimated the seroprevalence of SARS-CoV-2 in Sitakunda to have been 64.1% (95% CrI 60.0–68.1%) among all individuals and 63.4% (95% CrI 59.2-67.6%) when considering only unvaccinated individuals (Table 1, Supplemental Table 3). We estimated a 7% higher risk of being seropositive in men compared to women (95% CrI: 1-13%). Risk generally increased with age, with those less than 10 years old having ≥33% lower risk than other age groups (Table 1, Supplemental Table 4). We found a similar adjusted seroprevalence in the population recruited before the lockdown (63.1%, 95% CrI 56.2-69.8%; n=665) and those collected after the lockdown (65.3%, 95% CrI 60.6-69.9%; n=1,643).

**Table 1.** Estimated seroprevalence of SARS-CoV-2 in Sitakunda Upazila adjusted for sex, age, household clustering, and test performance among all individuals, both vaccinated and unvaccinated.

| Variable | Category | Observations | Test positive | Test negative | Adjusted Seroprevalence (95% CrI) | Adjusted Relative risk (95% CrI) |
| --- | --- | --- | --- | --- | --- | --- |
| Age (years) | 0–4 | 90 | 37 (41.1%) | 53 (58.9%) | 47.1 (37.0-57.3) | 0.66 (0.51-0.81) |
|  | 5–9 | 174 | 71 (40.8%) | 103 (59.2%) | 45.0 (37.1-52.9) | 0.63 (0.51-0.74) |
|  | 10–14 | 258 | 140 (54.3%) | 118 (45.7%) | 58.8 (52.0-65.3) | 0.83 (0.73-0.94) |
|  | 15–24 | 482 | 305 (63.3%) | 177 (36.7%) | 67.2 (61.7-72.6) | 0.96 (0.88-1.05) |
|  | 25–34 | 381 | 258 (67.7%) | 123 (32.3%) | 69.7 (64.5-75.0) | reference |
|  | 35–44 | 325 | 225 (69.2%) | 100 (30.8%) | 74.0 (68.3-79.5) | 1.07 (0.97-1.17) |
|  | 45–54 | 250 | 180 (72.0%) | 70 (28.0%) | 73.8 (67.2-80.3) | 1.06 (0.96-1.17) |
|  | 55–64 | 208 | 132 (63.5%) | 76 (36.5%) | 69.0 (62.1-75.8) | 0.99 (0.88-1.10) |
|  | 65+ | 139 | 95 (68.3%) | 44 (31.7%) | 73.6 (65.8-81.1) | 1.06 (0.94-1.19) |
| Sex | Male | 1 072 | 690 (64.4%) | 382 (35.6%) | 66.7 (62.2-71.3) | 1.07 (1.02-1.13) |
|  | Female | 1 235 | 753 (61.0%) | 482 (39.0%) | 61.3 (56.9-65.6) | reference |
| Overall |  | 2 307 | 1443 (62.5%) | 864 (37.5%) | 64.1 (60.0-68.1) | N/A |

In the catchment area of this serosurvey, only one healthcare facility provided SARS-CoV-2 testing (Bangladesh Institute of Tropical and Infectious Diseases). Among the 2,400 individuals that were tested from April 2020 until May 31, 2021, 705 (29%) tested positive for SARS-CoV-2 by RT-PCR. Crudely extrapolating our serologic estimates, by multiplying the estimated population size by adjusted seroprevalence among unvaccinated, suggests that more than 200,000 infections occurred during the same period, a much higher detected case to infection ratio than has been documented in most settings across the world (*5-6*). However, though the infection to case ratio is high, a substantial portion of missed infections were likely symptomatic; among the 161 respondents who reported having any COVID-19 like symptoms, only 57% (n=92) said they actively sought healthcare for treatment and only 1% (n=15/1442) of those who were seropositive were ever tested.

## Conclusions

These results illustrate that prior to the June 2021 surge in cases in Bangladesh fueled by the Delta-variant, the majority of the population in Sitakunda had already been infected despite a relatively low incidence of reported virologically confirmed SARS-CoV-2 infections. These data also provide reassuring evidence on the immunogenicity of the Covishield/ChAdOx1 vaccine in this population, where almost all self-reported vaccinees were seropositive. Key limitations to these results include the relatively small geographic area covered by the survey and that we only assessed circulating antibodies to a single SARS-CoV-2 epitope, which does not fully capture the immune profile of individuals.

In Bangladesh, where cases captured by surveillance are limited by healthcare seeking, even in population dense settings, representative seroprevalence surveys can help us continue to track the evolution of this pandemic. In addition to providing important validation data on a widely used immunoassay, our results can help lay the foundation for understanding the role of variant strains on key epidemiologic parameters, including our understanding of re-infection, and help set expectations for SARS-CoV-2 control in the months to come in the study area and beyond.

## Data Availability

Data supporting these estimates are publicly available in the sitakunda-sarscov2-round1 repository in Github.

https://github.com/HopkinsIDD/sitakunda-sarscov2-round1

## Acknowledgements

This work was supported by Bill and Melinda Gates Foundation [grant number INV-021879]. The authors would like to thank the Ministry of Health and Family Welfare (MOHFW) of Bangladesh. The authors would also like to express their sincere thanks to the staff members of icddr,b for their dedicated work in the field and laboratory during this pandemic situation. icddr,b is thankful and supported by the Governments of Bangladesh, Canada, Sweden, and the UK.

## Conflict of Interest

We declare no competing interests.

## Author Bio

Dr. Bhuiyan is an Associate Scientist at the International Centre for Diarrhoeal Disease Research, Bangladesh. His research interests include immunology, vaccinology, and enteric infections.

---

**Supplemental Table 1.**
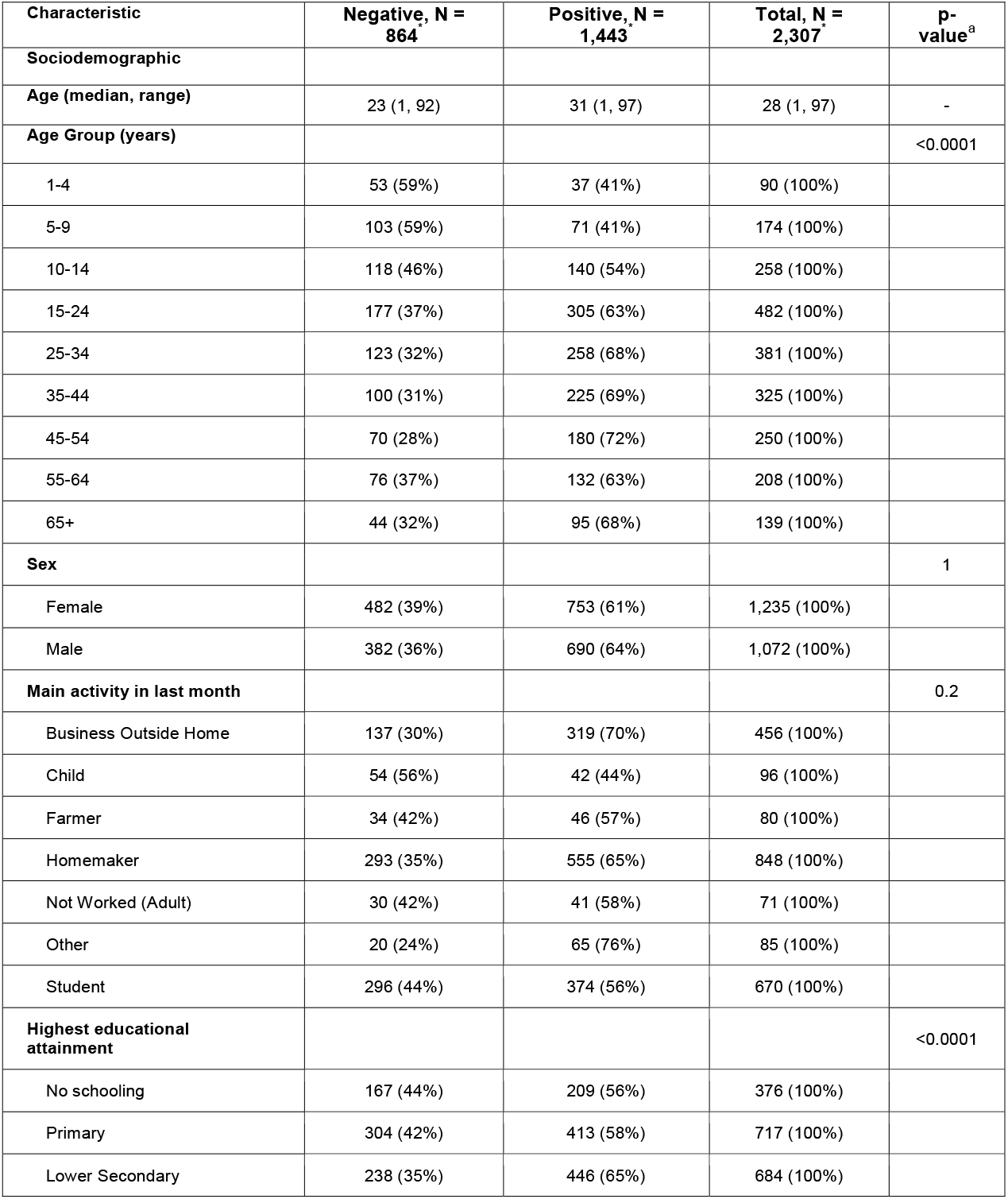
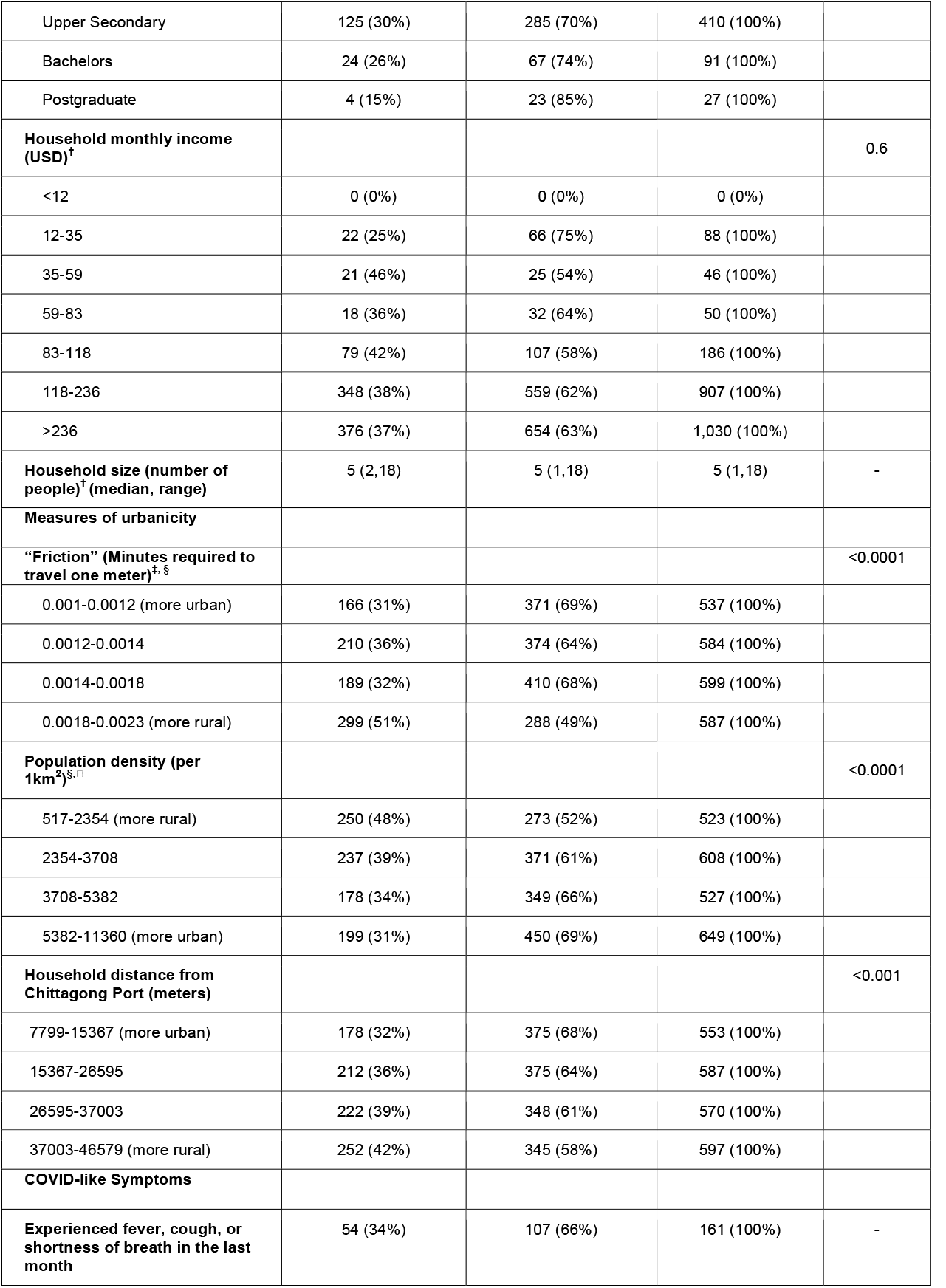
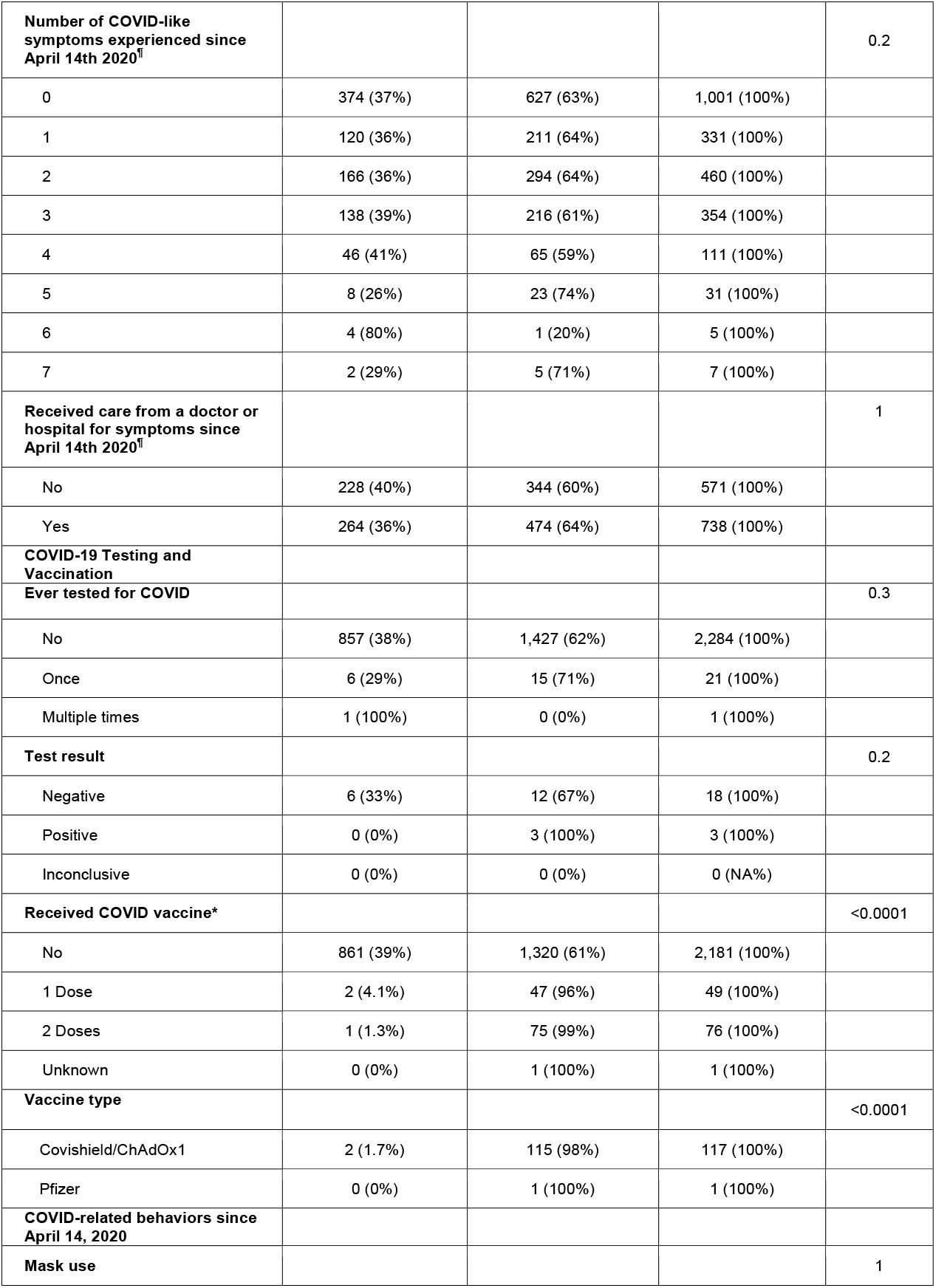
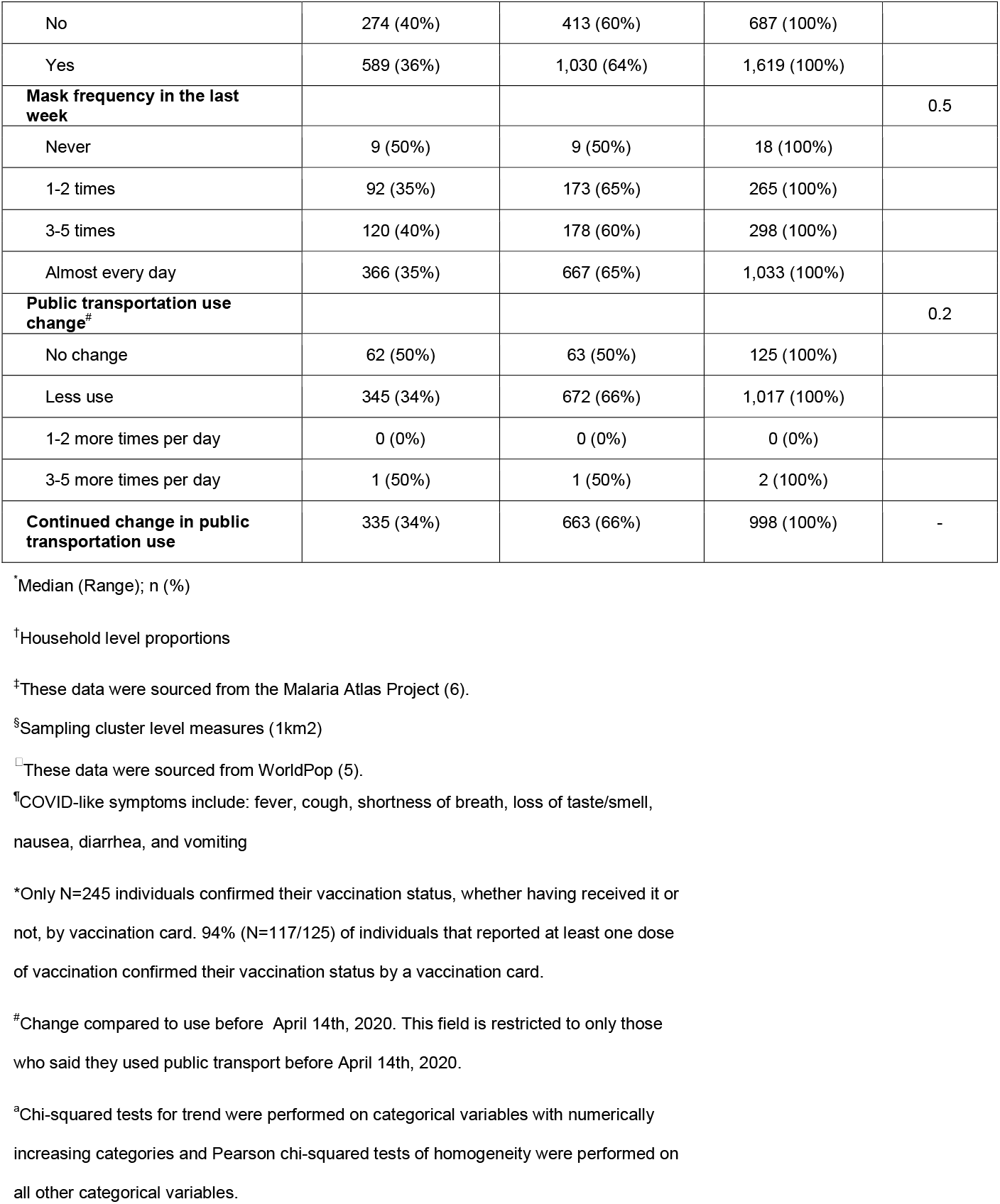
Descriptive statistics for serosurvey participants (n=2,307) in Sitakunda Upazila by seropositivity. This table includes sociodemographic factors, measures of urbanicity, COVID-like symptoms, COVID testing and vaccination, and COVID-related behaviors.

**Supplemental Table 2.** Descriptive statistics for unvaccinated, serosurvey participants (n=2,181) by seropositivity in Sitakunda Upazila. This table includes sociodemographic factors, measures of urbanicity, COVID-like symptoms, COVID testing and vaccination, and COVID-related behaviors.

| Characteristic | Negative, N = 268<sup>*</sup> | Positive, N = 371<sup>*</sup> | Total, N = 2,181<sup>*</sup> | p-value<sup>a</sup> |
| --- | --- | --- | --- | --- |
| **Sociodemographic** |  |  |  |  |
| **Age (median, range)** | 23 (1, 92) | 29 (1, 97) | 26 (1, 97) | - |
| **Age Group (years)** |  |  |  | <0.0001 |
| 1-4 | 53 (59%) | 37 (41%) | 90 (100%) |  |
| 5-9 | 103 (59%) | 71 (41%) | 174 (100%) |  |
| 10-14 | 118 (46%) | 140 (54%) | 258 (100%) |  |
| 15-24 | 176 (37%) | 303 (63%) | 479 (100%) |  |
| 25-34 | 123 (33%) | 249 (67%) | 372 (100%) |  |
| 35-44 | 100 (34%) | 198 (66%) | 298 (100%) |  |
| 45-54 | 69 (32%) | 147 (68%) | 216 (100%) |  |
| 55-64 | 76 (44%) | 97 (56%) | 173 (100%) |  |
| 65+ | 43 (36%) | 78 (64%) | 121 (100%) |  |
| **Sex** |  |  |  | 1 |
| Female | 480 (41%) | 700 (59%) | 1,180 (100%) |  |
| Male | 381 (38%) | 620 (62%) | 1,001 (100%) |  |
| **Main activity in last month** |  |  |  | 0.2 |
| Business Outside Home | 136 (32%) | 284 (68%) | 420 (100%) |  |
| Child | 54 (56%) | 42 (44%) | 96 (100%) |  |
| Farmer | 34 (45%) | 42 (55%) | 76 (100%) |  |
| Homemaker | 291 (37%) | 494 (63%) | 785 (100%) |  |
| Not Worked (Adult) | 30 (49%) | 31 (51%) | 61 (100%) |  |
| Other | 20 (27%) | 53 (73%) | 73 (100%) |  |
| Student | 296 (44%) | 373 (56%) | 669 (100%) |  |
| **Highest educational attainment** |  |  |  | <0.0001 |
| No schooling | 166 (45%) | 199 (55%) | 365 (100%) |  |
| Primary | 302 (44%) | 392 (56%) | 694 (100%) |  |
| Lower Secondary | 238 (37%) | 414 (63%) | 652 (100%) |  |
| Upper Secondary | 125 (34%) | 248 (66%) | 373 (100%) |  |
| Bachelors | 24 (32%) | 51 (68%) | 75 (100%) |  |
| Postgraduate | 4 (20%) | 16 (80%) | 20 (100%) |  |
| **Household monthly income (USD)<sup>†</sup>** |  |  |  | 0.5 |
| <12 | 0 (0%) | 0 (0%) | 0 (0%) |  |
| 12-35 | 22 (28%) | 58 (72%) | 80 (100%) |  |
| 35-59 | 21 (49%) | 22 (51%) | 43 (100%) |  |
| 59-83 | 18 (36%) | 32 (64%) | 50 (100%) |  |
| 83-118 | 78 (43%) | 102 (57%) | 180 (100%) |  |
| 118-236 | 347 (40%) | 517 (60%) | 864 (100%) |  |
| >236 | 375 (39%) | 589 (61%) | 964 (100%) |  |
| **Household size (number of people)<sup>†</sup> (median, range)** | 5 (2,18) | 6 (1,18) | 5 (1,18) | - |
| **Measures of urbanicity** |  |  |  |  |
| **“Friction” (Minutes required to travel one meter)<sup>†, §</sup>** |  |  |  | <0.0001 |
| 0.001-0.0012 (more urban) | 165 (33%) | 331 (67%) | 496 (100%) |  |
| 0.0012-0.0014 | 209 (38%) | 348 (62%) | 557 (100%) |  |
| 0.0014-0.0018 | 189 (34%) | 375 (66%) | 564 (100%) |  |
| 0.0018-0.0023 (more rural) | 298 (53%) | 266 (47%) | 564 (100%) |  |
| **Population density (per 1km<sup>2</sup>)<sup>§, ¶</sup>** |  |  |  | <0.0001 |
| 517-2354 (more rural) | 249 (50%) | 250 (50%) | 499 (100%) |  |
| 2354-3708 | 236 (40%) | 347 (60%) | 583 (100%) |  |
| 3708-5382 | 178 (36%) | 313 (64%) | 491 (100%) |  |
| 5382-11360 (more urban) | 198 (33%) | 410 (67%) | 608 (100%) |  |
| **Household distance from Chittagong Port (meters)** |  |  |  | <0.001 |
| 7799-15367 (more urban) | 177 (35%) | 336 (65%) | 513 (100%) |  |
| 15367-26595 | 212 (37%) | 355 (63%) | 567 (100%) |  |
| 26595-37003 | 221 (40%) | 325 (60%) | 546 (100%) |  |
| 37003-46579 (more rural) | 251 (45%) | 304 (55%) | 555 (100%) |  |
| **COVID-like Symptoms** |  |  |  |  |
| **Experienced fever, cough, or shortness of breath in the last month** | 54 (36%) | 97 (64%) | 151 (100%) | - |
| **Number of COVID-like symptoms experienced since April 14th 2020<sup>†</sup>** |  |  |  | 0.4 |
| 0 | 372 (39%) | 570 (61%) | 942 (100%) |  |
| 1 | 120 (38%) | 197 (62%) | 317 (100%) |  |
| 2 | 166 (38%) | 266 (62%) | 432 (100%) |  |
| 3 | 138 (41%) | 200 (59%) | 338 (100%) |  |
| 4 | 46 (43%) | 60 (57%) | 106 (100%) |  |
| 5 | 8 (28%) | 21 (72%) | 29 (100%) |  |
| 6 | 3 (75%) | 1 (25%) | 4 (100%) |  |
| 7 | 2 (33%) | 4 (67%) | 6 (100%) |  |
| **Received care from a doctor or hospital for symptoms since April 14th 2020<sup>†</sup>** |  |  |  | 1 |
| No | 228 (41%) | 321 (59%) | 549 (100%) |  |
| Yes | 263 (38%) | 432 (62%) | 695 (100%) |  |
| **COVID-19 Testing** |  |  |  |  |
| **Ever tested for COVID** |  |  |  | 0.5 |
| No | 854 (40%) | 1,308 (60%) | 2,162 (100%) |  |
| Once | 6 (35%) | 11 (65%) | 17 (100%) |  |
| Multiple times | 1 (100%) | 0 (0%) | 1 (100%) |  |
| **Test result** |  |  |  | 0.2 |
| Negative | 6 (43%) | 8 (57%) | 14 (100%) |  |
| Positive | 0 (0%) | 3 (100%) | 3 (100%) |  |
| Inconclusive | 0 (0%) | 0 (0%) | 0 (0%) |  |
| **COVID-related behaviors since April 14, 2020** |  |  |  |  |
| **Mask use** |  |  |  | 1 |
| No | 272 (41%) | 385 (59%) | 657 (100%) |  |
| Yes | 588 (39%) | 935 (61%) | 1,523 (100%) |  |
| **Mask frequency in the last week** |  |  |  | 0.9 |
| Never | 9 (53%) | 8 (47%) | 17 (100%) |  |
| 1-2 times | 92 (36%) | 163 (64%) | 255 (100%) |  |
| 3-5 times | 120 (41%) | 170 (59%) | 290 (100%) |  |
| Almost every day | 365 (38%) | 591 (62%) | 956 (100%) |  |
| **Public transportation use<sup>#</sup>** |  |  |  | 0.2 |
| No change | 61 (51%) | 58 (49%) | 119 (100%) |  |
| Less use | 344 (37%) | 598 (63%) | 942 (100%) |  |
| 1-2 more times per day | 0 (0%) | 0 (0%) | 0 (0%) |  |
| 3-5 more times per day | 1 (50%) | 1 (50%) | 2 (100%) |  |
| **Continued change in public transportation use** | 334 (36%) | 590 (64%) | 924 (100%) | - |

\* Median (Range); n (%)
† Household level proportions
‡ These data were sourced from the Malaria Atlas Project (6).
§ Sampling cluster level measures (1km<sup>2</sup>)
□ These data were sourced from WorldPop (5).
¶ COVID-like symptoms include: fever, cough, shortness of breath, loss of taste/smell, nausea, diarrhea, and vomiting
<sup>#</sup> Change compared to use before April 14th, 2020. This field is restricted to only those who said they used public transport before April 14th, 2020.
<sup>a</sup> Chi-squared tests for trend were performed on categorical variables with numerically increasing categories and Pearson chi-squared tests of homogeneity were performed on all other categorical variables.

**Supplemental Table 3.**
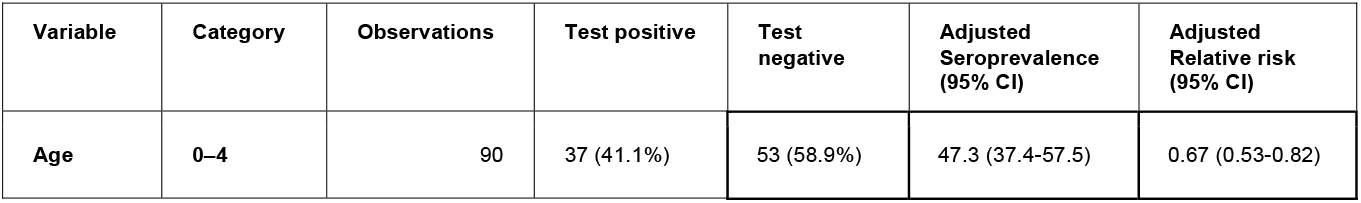
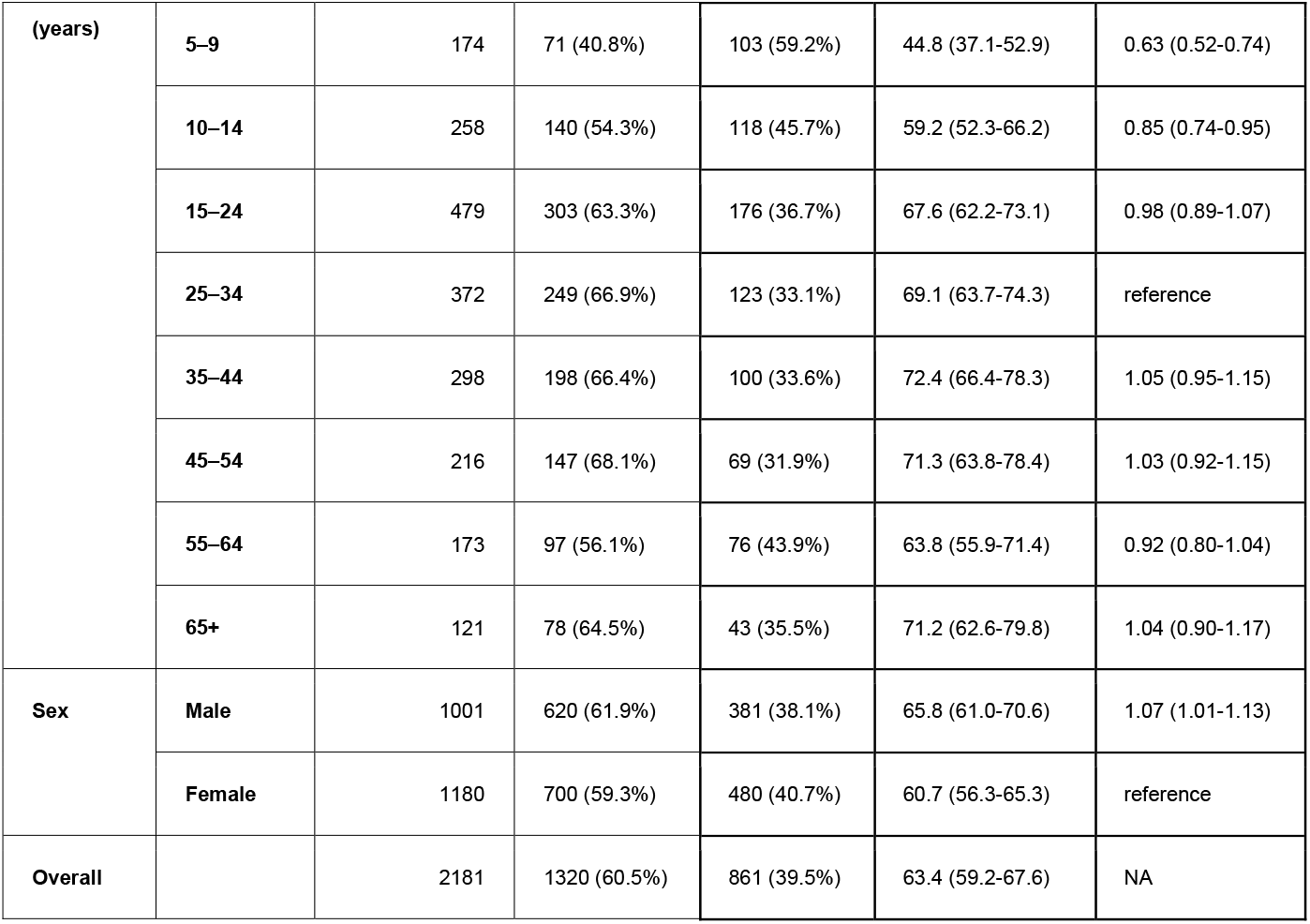
Estimated seroprevalence of SARS-CoV-2 in Sitakunda Upazila adjusted for sex, age, household clustering, and test performance among unvaccinated individuals.

| Variable | Category | Observations | Test positive | Test negative | Adjusted Seroprevalence (95% CI) | Adjusted Relative risk (95% CI) |
| --- | --- | --- | --- | --- | --- | --- |
| Age | 0–4 | 90 | 37 (41.1%) | 53 (58.9%) | 47.3 (37.4-57.5) | 0.67 (0.53-0.82) |
| **(years)** | **5–9** | 174 | 71 (40.8%) | 103 (59.2%) | 44.8 (37.1-52.9) | 0.63 (0.52-0.74) |
|  | **10–14** | 258 | 140 (54.3%) | 118 (45.7%) | 59.2 (52.3-66.2) | 0.85 (0.74-0.95) |
|  | **15–24** | 479 | 303 (63.3%) | 176 (36.7%) | 67.6 (62.2-73.1) | 0.98 (0.89-1.07) |
|  | **25–34** | 372 | 249 (66.9%) | 123 (33.1%) | 69.1 (63.7-74.3) | reference |
|  | **35–44** | 298 | 198 (66.4%) | 100 (33.6%) | 72.4 (66.4-78.3) | 1.05 (0.95-1.15) |
|  | **45–54** | 216 | 147 (68.1%) | 69 (31.9%) | 71.3 (63.8-78.4) | 1.03 (0.92-1.15) |
|  | **55–64** | 173 | 97 (56.1%) | 76 (43.9%) | 63.8 (55.9-71.4) | 0.92 (0.80-1.04) |
|  | **65+** | 121 | 78 (64.5%) | 43 (35.5%) | 71.2 (62.6-79.8) | 1.04 (0.90-1.17) |
| **Sex** | **Male** | 1001 | 620 (61.9%) | 381 (38.1%) | 65.8 (61.0-70.6) | 1.07 (1.01-1.13) |
|  | **Female** | 1180 | 700 (59.3%) | 480 (40.7%) | 60.7 (56.3-65.3) | reference |
| **Overall** |  | 2181 | 1320 (60.5%) | 861 (39.5%) | 63.4 (59.2-67.6) | NA |

**Supplementary Table 4.** The number of positive controls used to estimate the empirical sensitivity of the Wantai total Ab assay and SARS-CoV-2 positivity by time since symptom onset.

| **Time post-symptom onset** | **Number of samples** | **% Seropositive (n)** |
| --- | --- | --- |
| 3-13 days | 2 | 100% (2) |
| 14-30 days | 6 | 83.3% (5) |
| 31-60 days | 5 | 100% (5) |
| 61-90 days | 7 | 100% (7) |
| 91-120 days | 7 | 85.7% (6) |
| 121-150 days | 10 | 80% (8) |
| 151-180 days | 12 | 83.3% (10) |
| 181-210 days | 11 | 72.7% (8) |
| 211-240 days | 9 | 100% (9) |
| 241-275 days | 12 | 100% (12) |

**Supplemental Figure 1.** Map of the study population in the Sitakunda Upazila (in green) in the Chattogram District of Bangladesh. The 580 enrolled households sampled in the serosurvey by enrollment time (pre versus post lockdown) and the two healthcare facilities in Sitakunda (BITID and the Sitakunda UHC) are indicated on the right side of the figure.

**Supplemental Figure 2.** Map of the sampled clusters and sampled dwellings within clusters in the Sitakunda Upazila by samples drawn at three different periods: first draw (March 2021), second draw (May 2021), third draw (June 2021). Three separate sample draws were conducted due to interruption from the nationally imposed lockdown and a large percentage of non-residential structures among housing structures sampled from the satellite imagery. (A) Clusters were sampled 41 times with 14 structures each during the first two draws and 12 structures during the last draw. (B) We oversampled the number of structures by 40% to account for nonresidential buildings for a total of 574 sampled structures for the first two draws and 492 for the last draw. (C) Households were enrolled across the entire subdistrict of Sitakunda during each enrollment period and by sample draw (households enrolled pre-lockdown were only drawn from the first sample).

**Supplemental Figure 3.** Average seropositivity among all participants in each sampled cluster (1 km^2^) by (A) population density (B) the minutes required to travel one minute (shown with all cluster data points and excluding the cluster with an outlier value) and (C) average household distance to Chittagong Port, the center of the Chattogram City. Loess smoothing is shown in blue with 95% confidence intervals shown in grey.

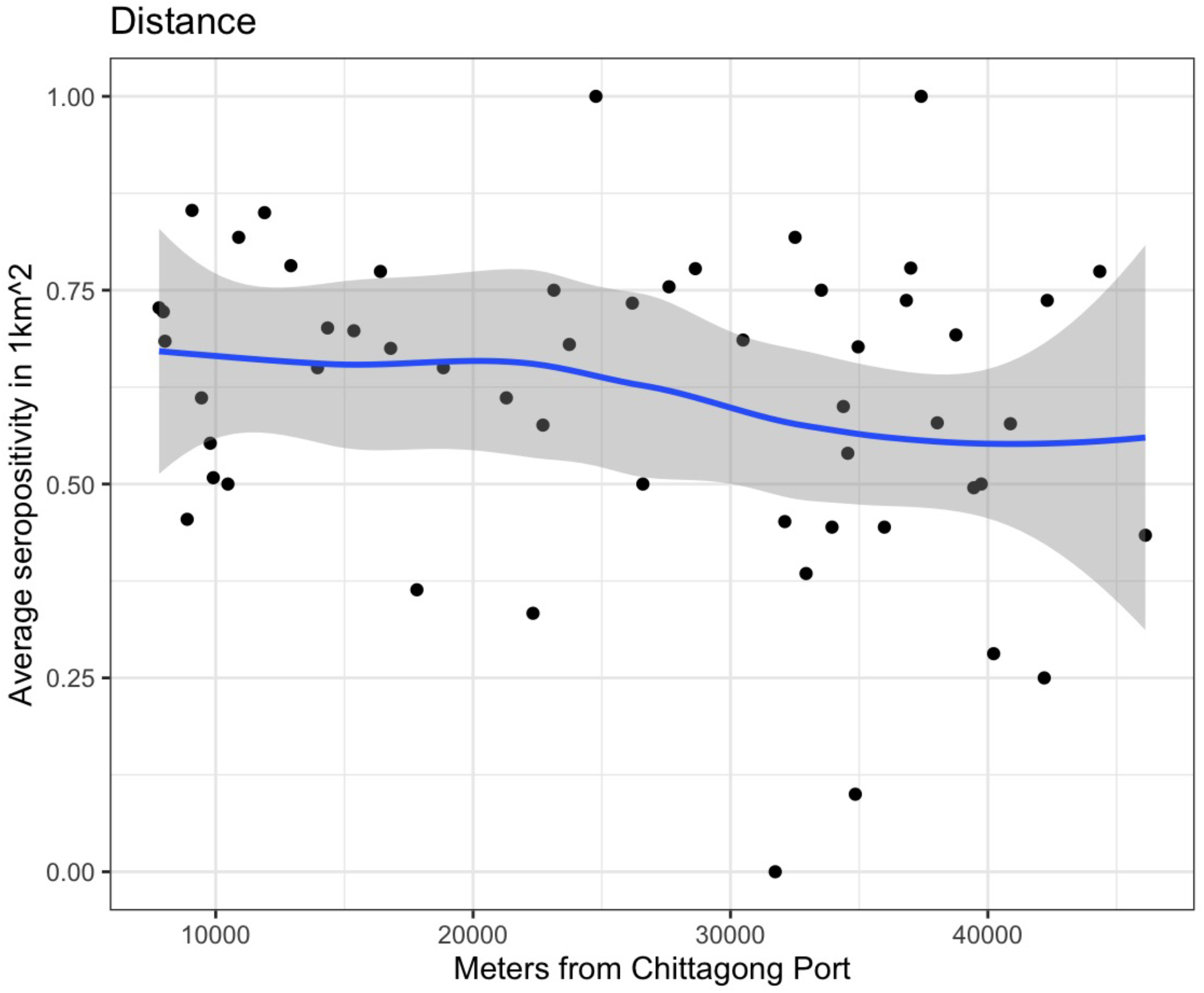

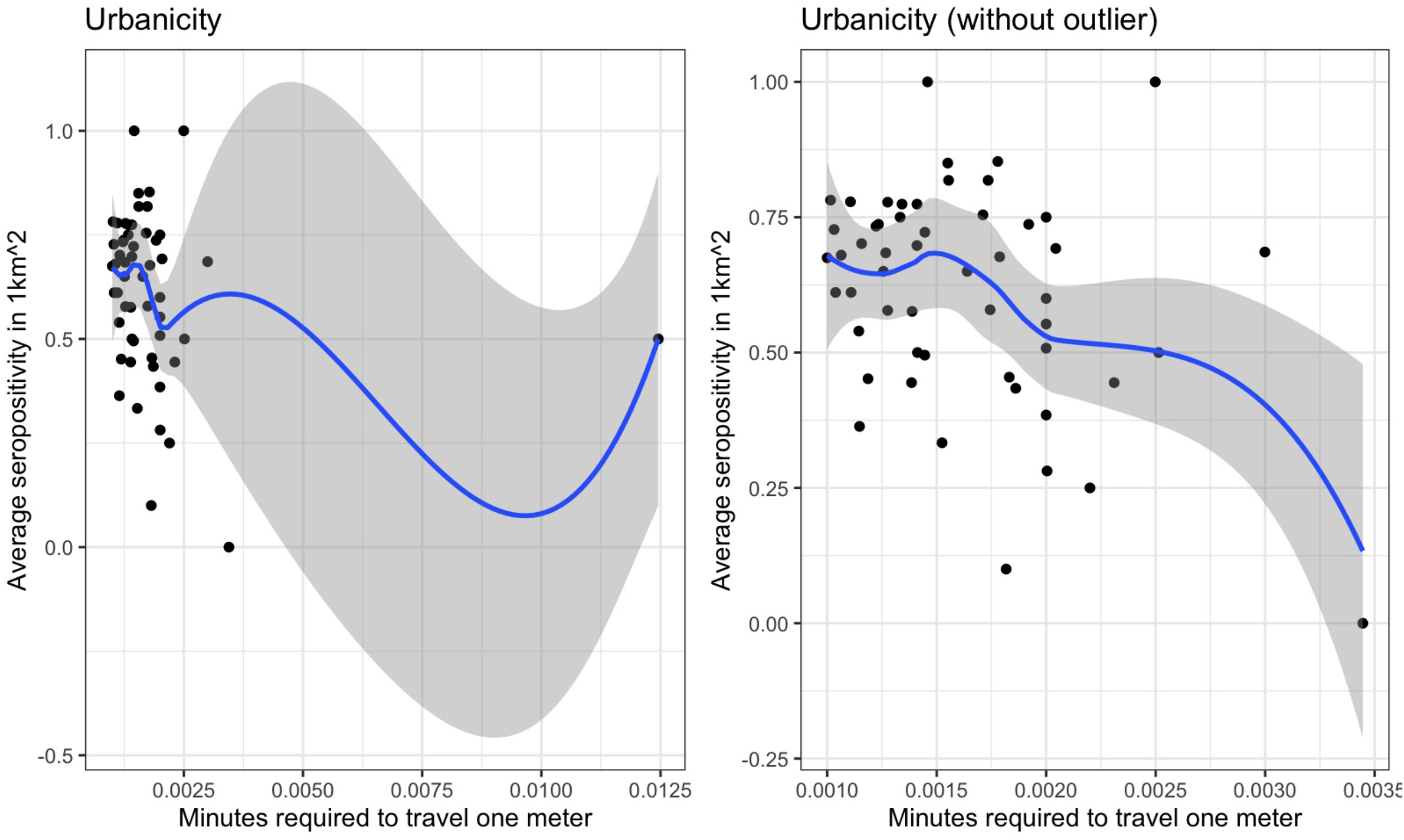

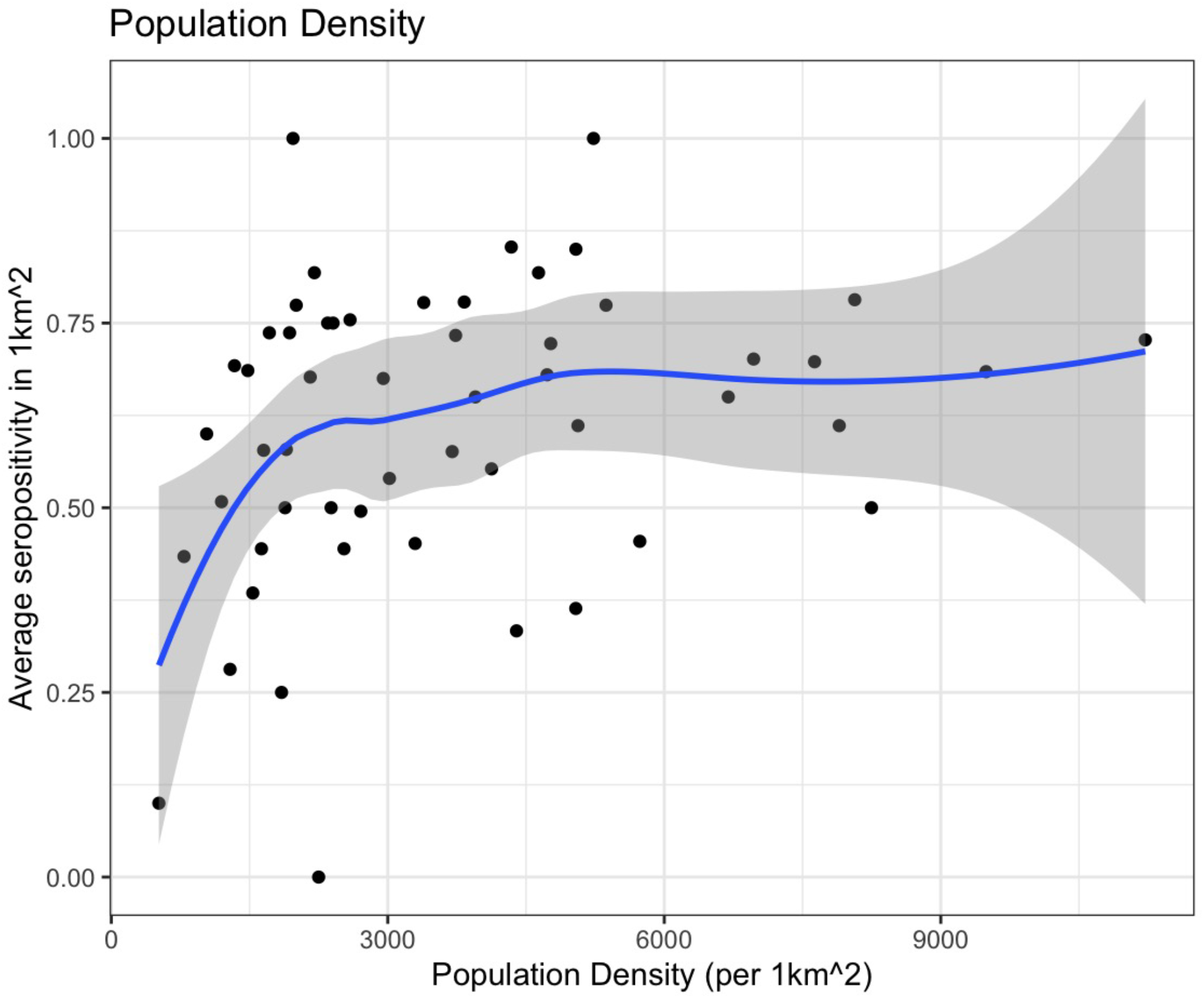

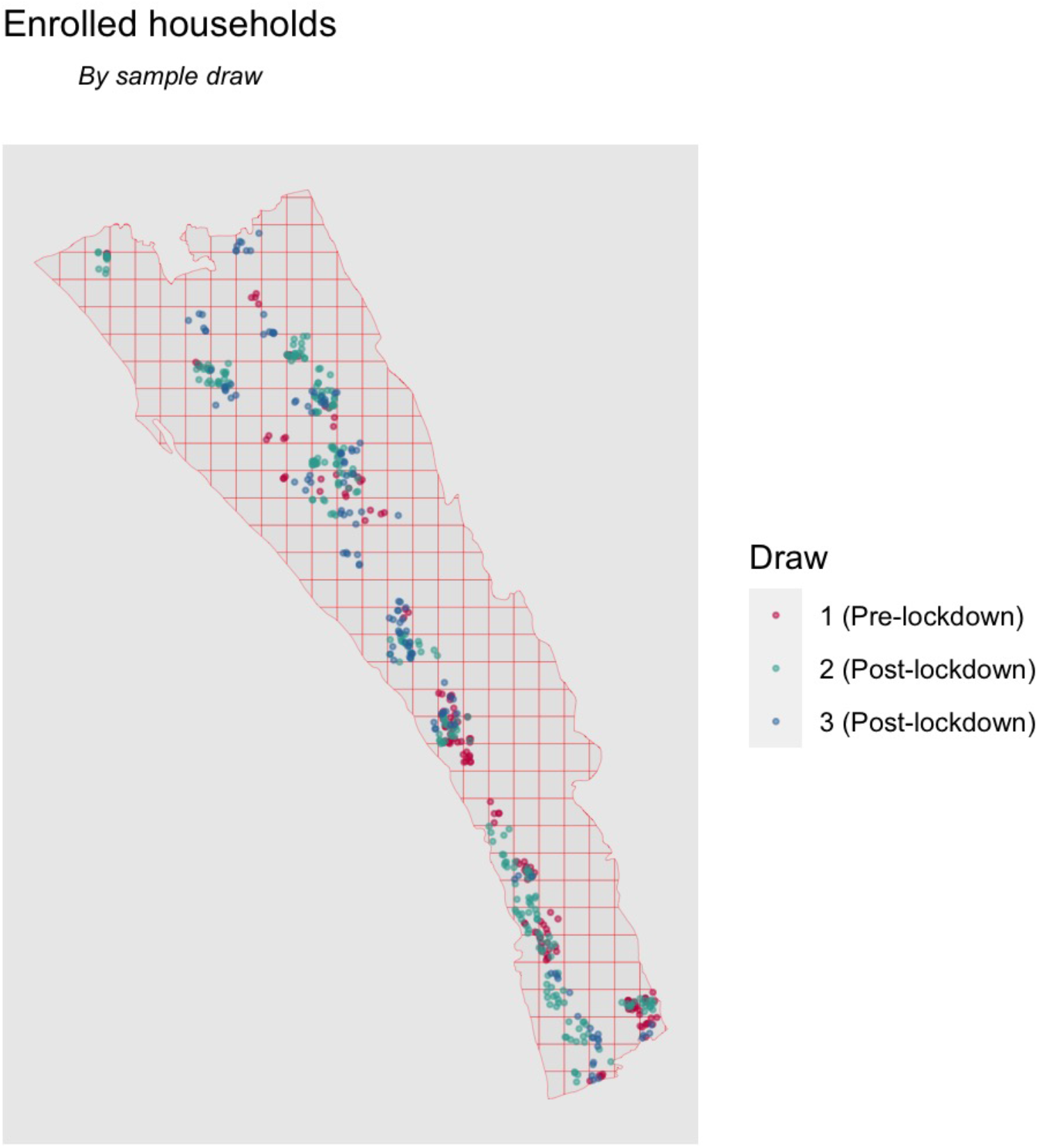

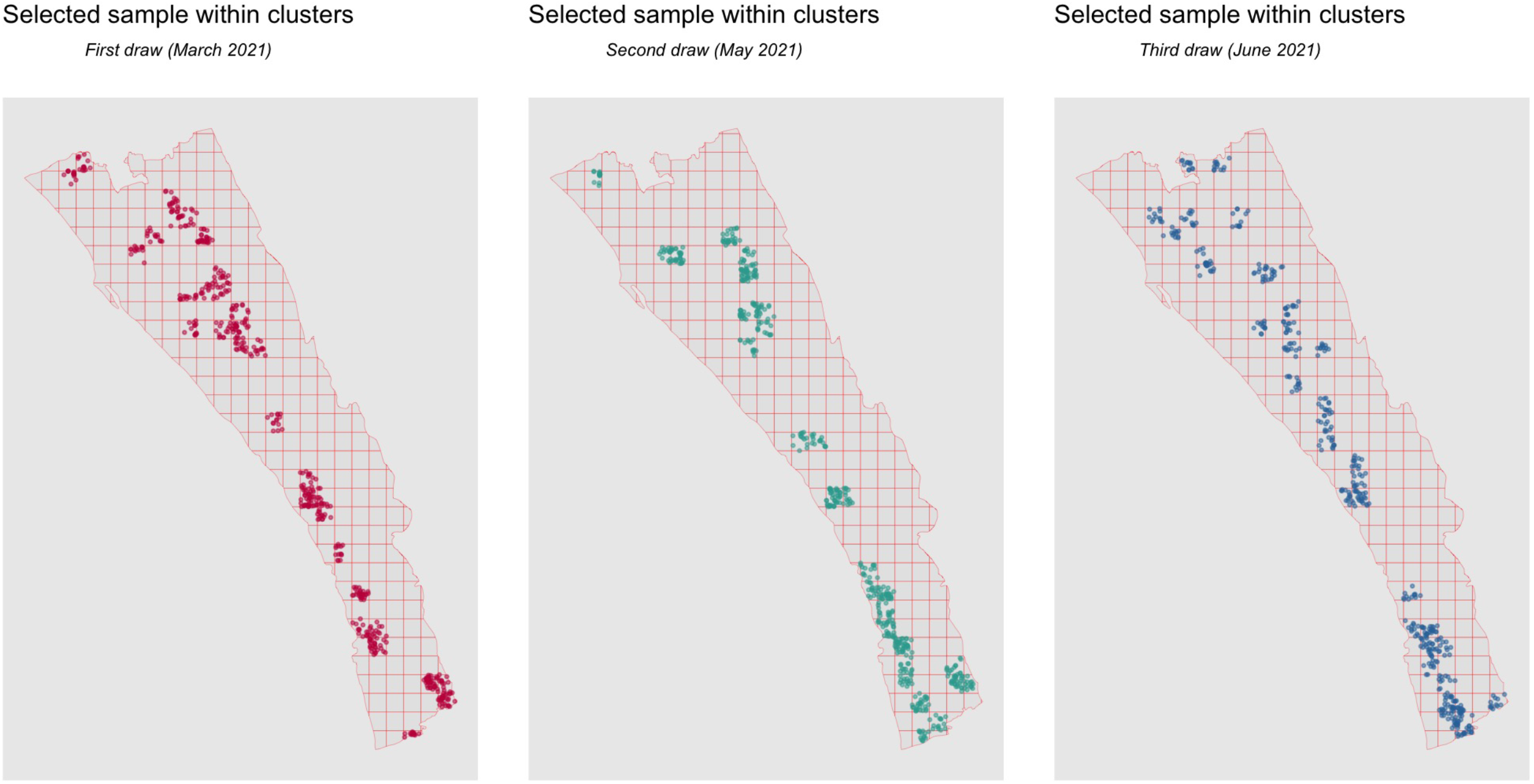

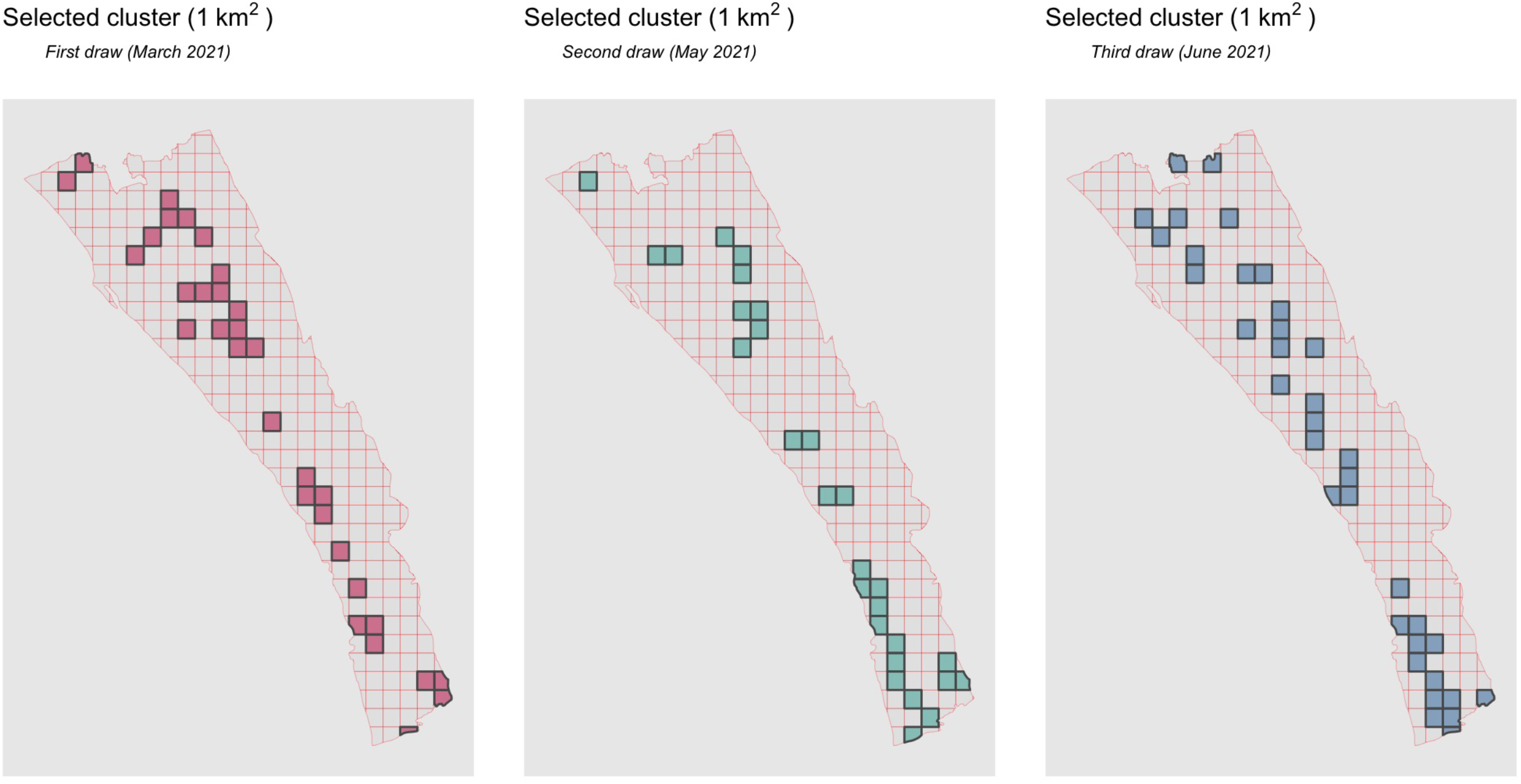

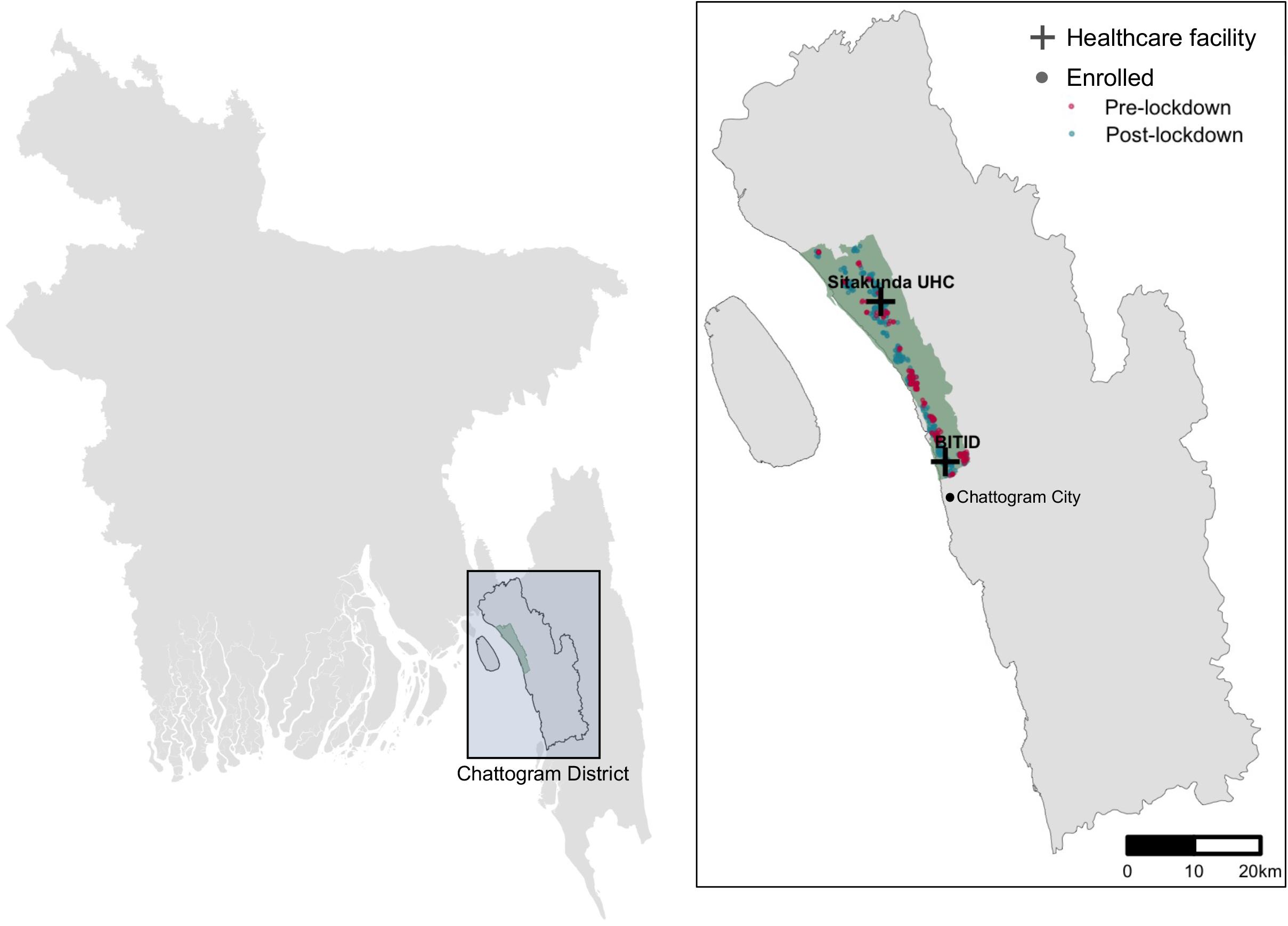

## Notes

### Competing Interest Statement

The authors have declared no competing interest.

### Author Declarations

IRB approval was given by ethical review committees from both Johns Hopkins University and International Centre for Diarrhoeal Disease Research, Bangladesh (icddr,b).

### Summary of Updates

Updated to include more data from after lockdown.

