## Supplement for "SARS-CoV-2 seroprevalence in Chattogram, Bangladesh before the Delta surge, March-June 2021"

*Supplemental Materials*

**Supplemental Methods**

*Study population*

The study population was the population of the Sitakunda Upazila (subdistrict) within the Chattogram district in Southeastern Bangladesh. This study was originally designed to estimate seroincidence of *Vibrio cholerae* infection through serial serosurveys. Participant enrollment was focused in a population that is known to primarily seek care for acute watery diarrhea at the Bangladesh Institute for Tropical and Infectious Diseases (BITID) and the Sitakunda Upazila Health Complex (UHC), two healthcare facilities in the Sitakunda subdistrict. This area was additionally selected to minimize loss to follow-up in subsequent survey rounds as it is less urban than Chattogram City (bordering the southern end of Sitakunda) and the population is less transient. Individuals living in Sitakunda have few options for accessing traditional healthcare facilities by road due to the road networks in the Chattogram district, except for BITID and the Sitakunda UHC (Supplemental Figure 1).

*Survey methods*

We conducted this household survey between March 26–April 13, 2021, before the lockdown in Bangladesh, and between May 23-June 13, 2021, after the lockdown was lifted. The sampling frame was constructed using satellite imagery of the Sitakunda subdistrict (collected in the fourth quarter of 2020) and applying a deep-learning algorithm to identify single- and multi-story structures, based on sizes of the structures and their shadows and validated by manual review using an external agency (*1*). Prior to the survey, a dwelling assessment was conducted on a random selection of satellite identified households to determine the proportion of identified dwellings that are residential and the distribution of the number of households in multistory residential structures.

Assuming a SARS-CoV-2 IgG seroprevalence of 25%, we estimated that a sample size of 1,632 individuals (~408 households) would lead to a 95% confidence interval half-width of 1.8 percentage points. This assumed a design effect of 3, given a household-level ICC of 0.676 and an average household size of 4 (*2*).

We used a two-stage cluster sampling approach to select 574 structures identified from the satellite imagery. We oversampled the number of structures to account for non-residential structures (~40%). The Sitakunda subdistrict was divided into clusters one square kilometer in size, and 41 clusters with 14 structures each were randomly selected with replacement (32 unique clusters) (Supplemental Figure 2). Weights accounted for the estimated number of households in multistory units using data from the dwelling assessment of the area. Data collection efforts were terminated after visiting 251 structures and enrolling at least one household in 78% (n=25/32) of clusters due to the national lockdown imposed on April 5^th^, 2021 in response to the rising number of COVID-19 cases.

To reach the intended sample size of 1,632 individuals at a single time point, after the imposed lockdown, a new two-stage cluster sample was drawn and 574 structures, excluding the 251 already visited structures, were selected. We oversampled the number of structures to again account for non-residential structures; 41 clusters with 14 structures each were randomly selected with replacement (28 unique clusters). All 574 structures were visited before the sample size was reached resulting in a third draw of the two-stage cluster sample. Therefore, a third sample draw of 492 new structures excluding the previously visited structures (251 + 574 structures) was conducted; 41 clusters with 12 structures each were randomly selected with replacement (36 unique clusters). When a structure included multiple households, the teams enumerated the number of households and used a random number generator on their data collection tablets to select one household at random.

During each household visit, study staff explained the study to the head of each household (or designee) and first asked for verbal consent from the household head to discuss the study with other members of the household. Household members were eligible if they were at least one years old, if they had resided in that village for the past year, intended to continue living there for the next year, and did not plan to spend more than two consecutive months away from the village in the coming year. Each eligible individual was asked for written informed consent to participate if she/he was home during one of up to three repeated visits. This study was approved by the icddr,b Research and Ethics Review Committees and the Johns Hopkins Bloomberg School of Public Health Institutional Review Board.

*Analysis*

We estimated seroprevalence using a Bayesian regression model with fixed effects for age group and sex and a random effect for household clustering and adjusting for immunoassay performance (*2*). Within this model we also estimate and correct for the immunoassay performance (sensitivity and specificity) using data collected in Dhaka from positive and negative controls. We sampled from the posterior distribution using Hamiltonian MCMC as implemented in Stan (used through rstan 2.21.1) and ran four separate chains for 1,500 iterations each (*3*). We used the R-hat statistic to evaluate convergence. We post-stratified model results using 2011 Census data on the age and sex disaggregation of the Sitakunda Upazila population (*4*).

Measures of urbanicity, such as population density (using the 2015 population estimates from WorldPop (*5*)) and time required to travel one meter for each enrolled cluster (using the 2019 friction surface enumerating land-based travel speed with access to motorized transport from Malaria Atlas Project (*6*)), and household distance to Chittagong Port, the center of Chattogram City, for each enrolled cluster were categorized by quartiles and assigned to participants. We used a chi-squared test for trend to determine whether seropositivity was associated with increased levels of urbanicity (by various metrics). We also evaluated differences in average seropositivity in each enrolled cluster (1 km^2^) by population density, the minutes required to travel one minute, and the average household distance to Chittagong Port. Other variables with numerically increasing categories were also tested using a chi-squared test for trend in proportions to detect a relationship to seropositivity. All other categorical variables were tested using a Pearson chi-squared test for homogeneity (Supplementary Table 1 & Supplementary Table 2).

All analyses were performed in R. Data and source code to reproduce analyses are available at https://github.com/HopkinsIDD/sitakunda-sarscov2-round1.

**Supplemental Results**

*Study participation rates*

Among the 251 structures we visited from March 27 to April 13 (pre-lockdown), 170 (68%) were residential structures. We enrolled 99.4% (n=169/170) of the residential structures visited and later identified one enrolled household that consisted of two separate residences (i.e., two separate cooking pots and areas where individuals regularly sleep); 170 households were enrolled prior to the lockdown. We enrolled 72.6% of eligible household members (≥1 year of age) (n=667/919) and of the 667 individuals enrolled, 665 (99.7%) provided a blood sample.

After the lockdown, we visited a total of 841 structures and 561 (67%) were residential structures. We enrolled 410 (73%) of the residential structures and 76.7% (n=1672/2179) of age eligible household members into the study. Among the 1,672 individuals enrolled, 1,643 provided a blood sample. In total, 2,337 individuals (580 households) were enrolled in this study and 2,307 individual samples were used in this analysis.

**Supplemental Figure 1.**

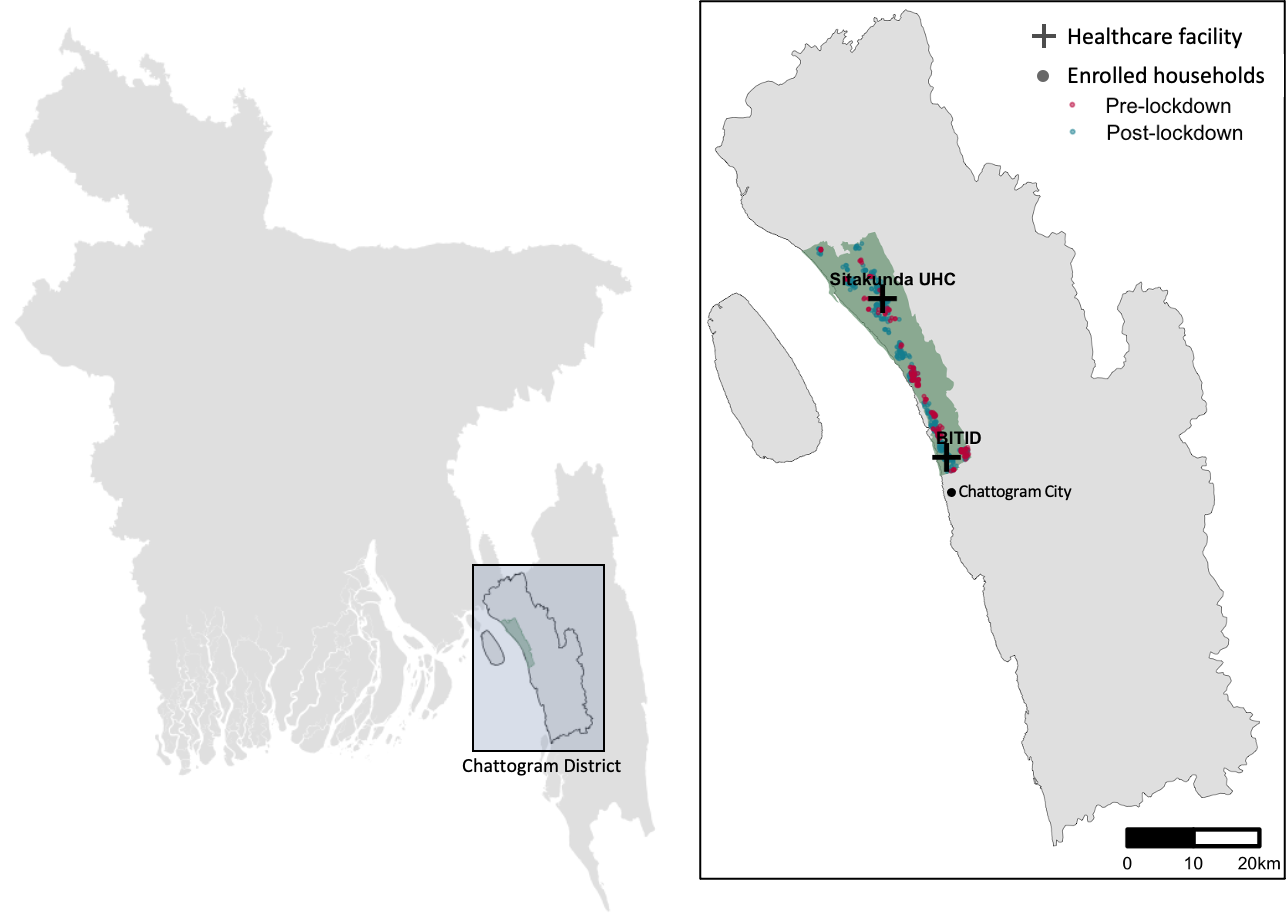

**Supplemental Figure 2. (A)**

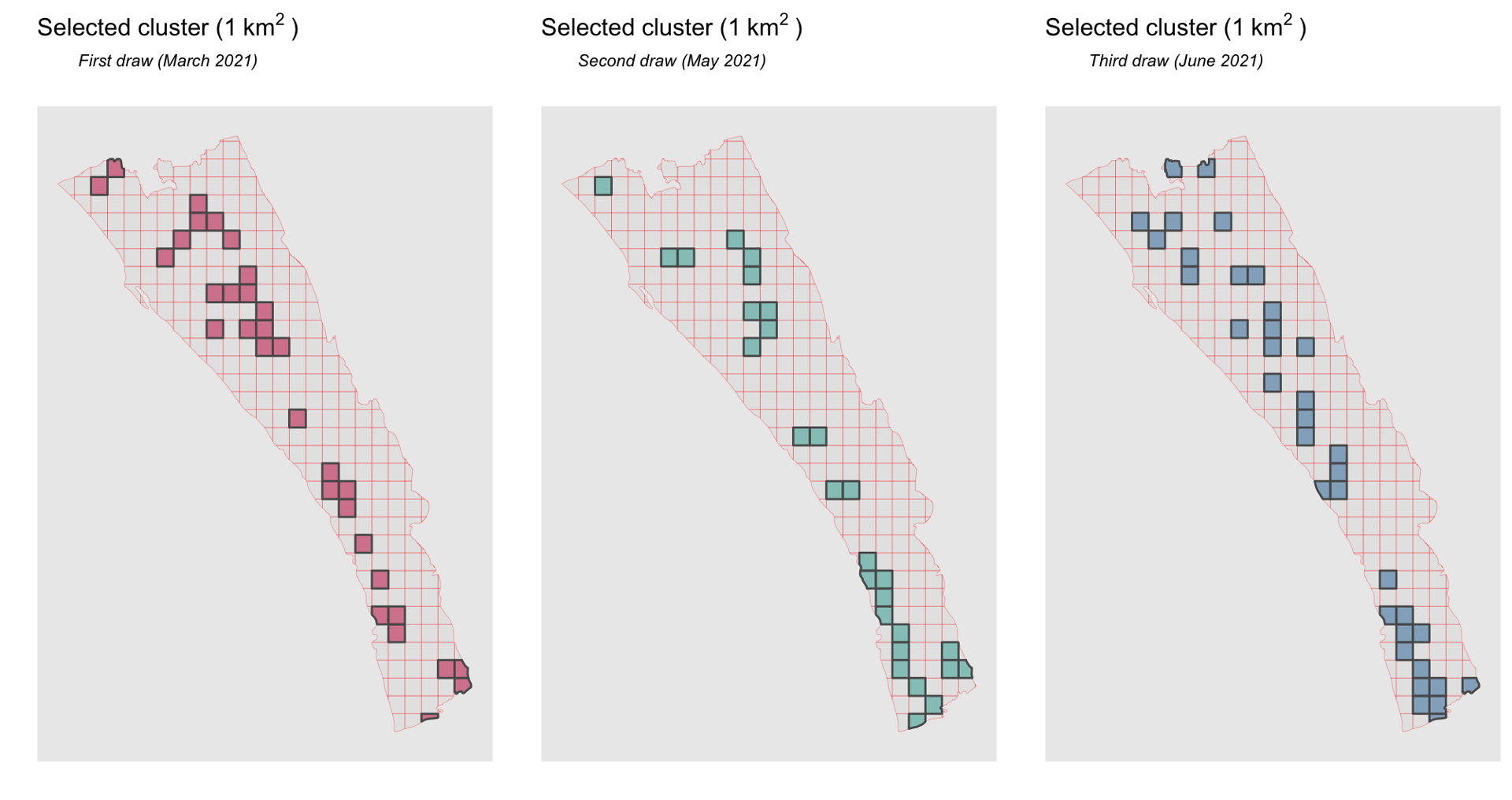

**Supplemental Figure 2. (B)**
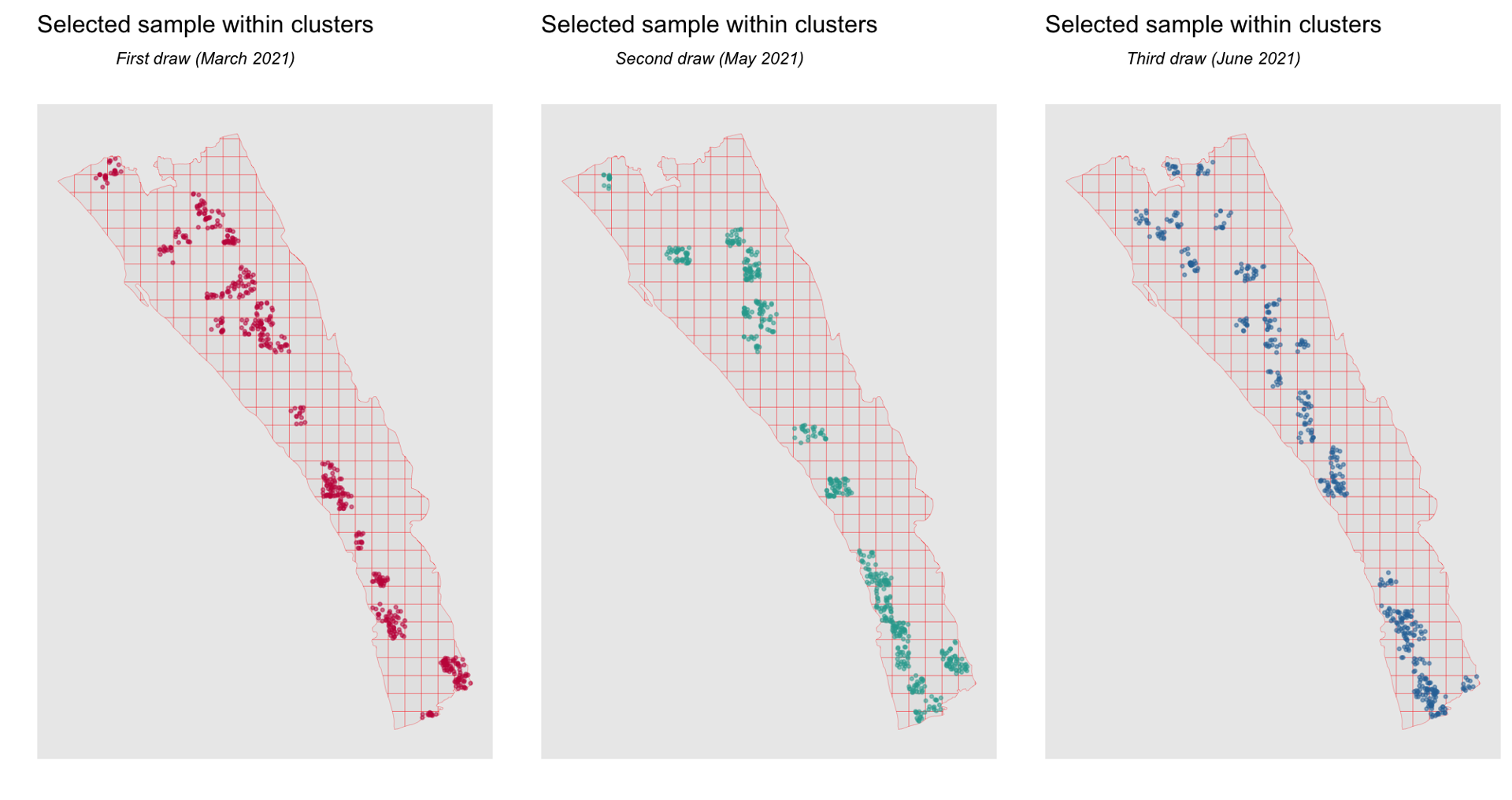

**Supplemental Figure 2. (C)**
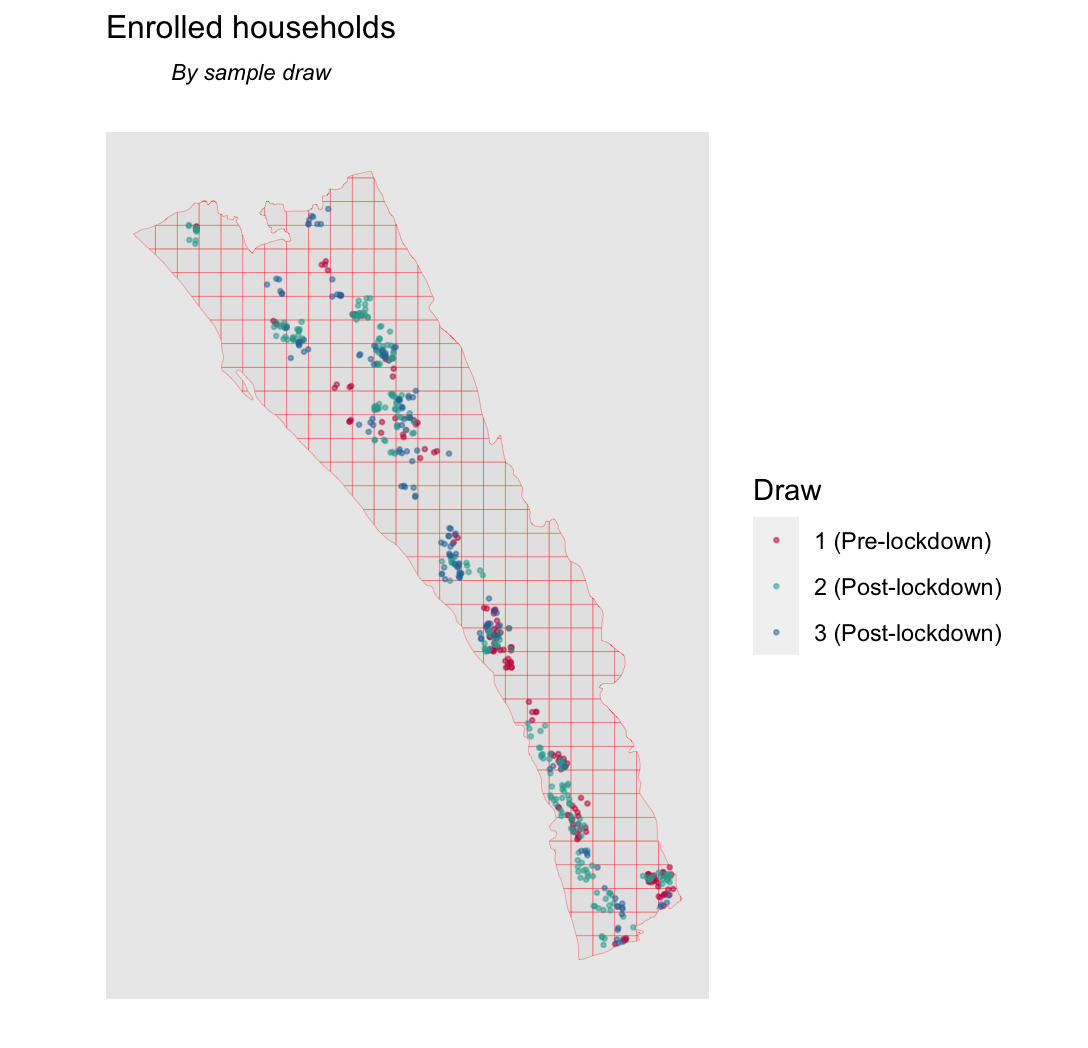

**Supplemental Figure 3. (A)**
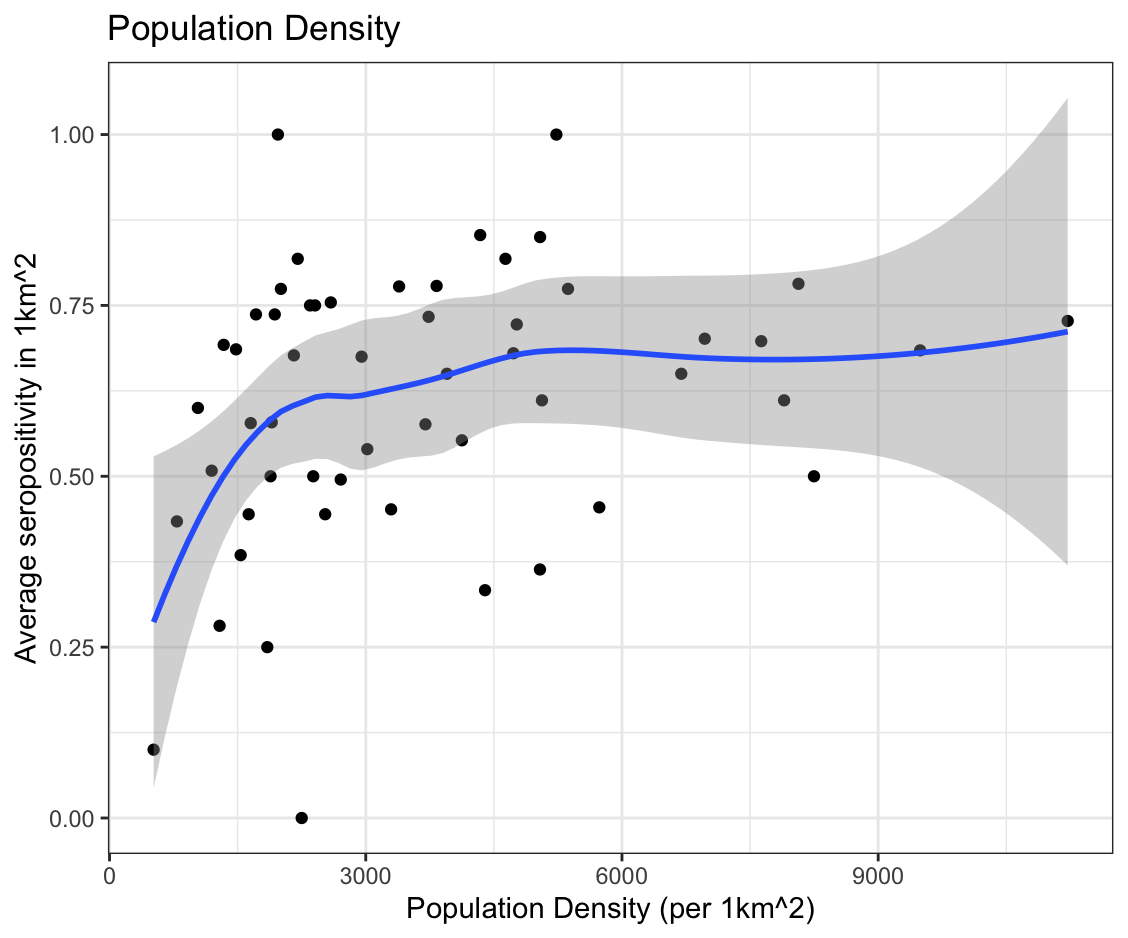

**Supplemental Figure 3. (B)**
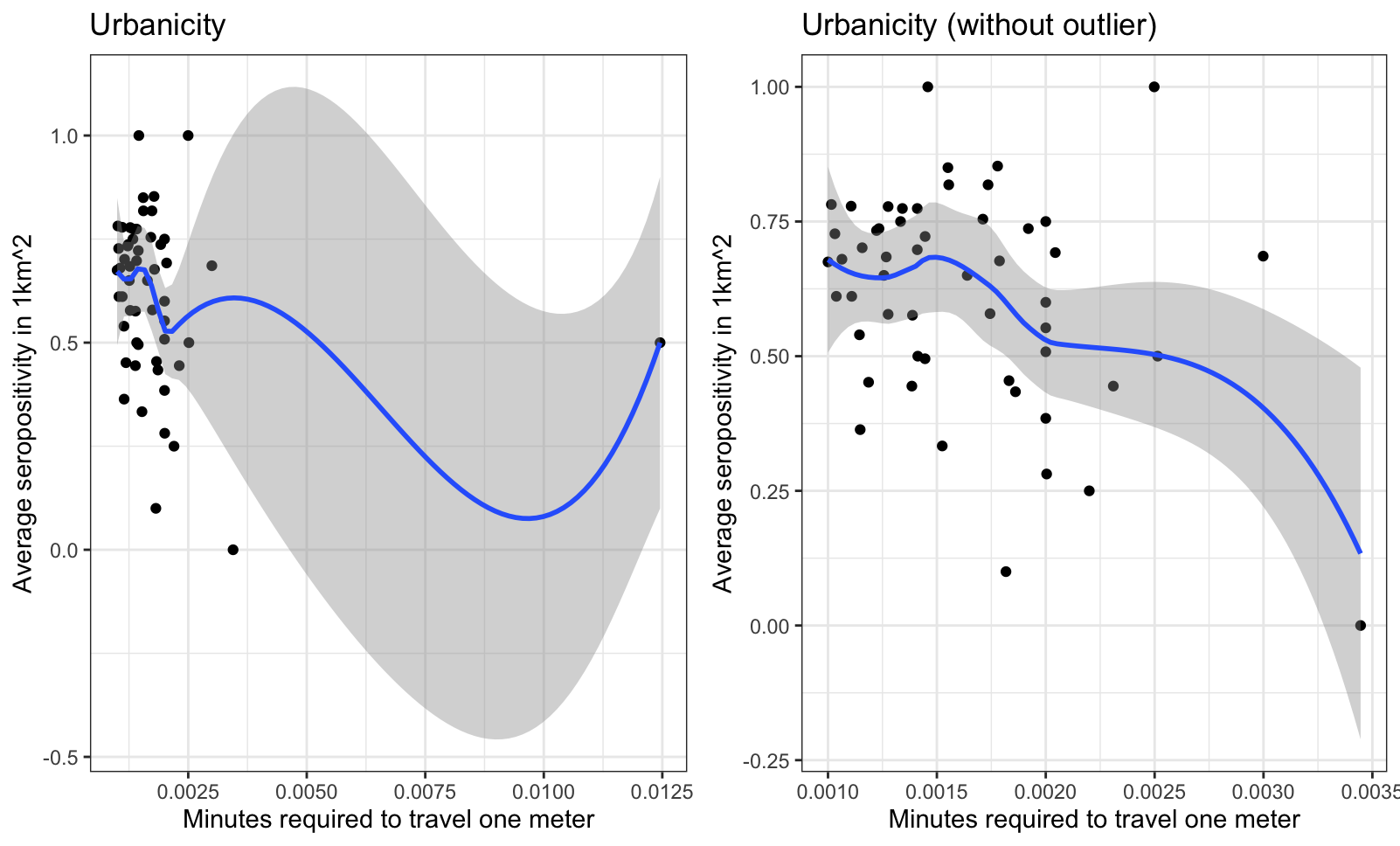

**Supplemental Figure 3. (C)**
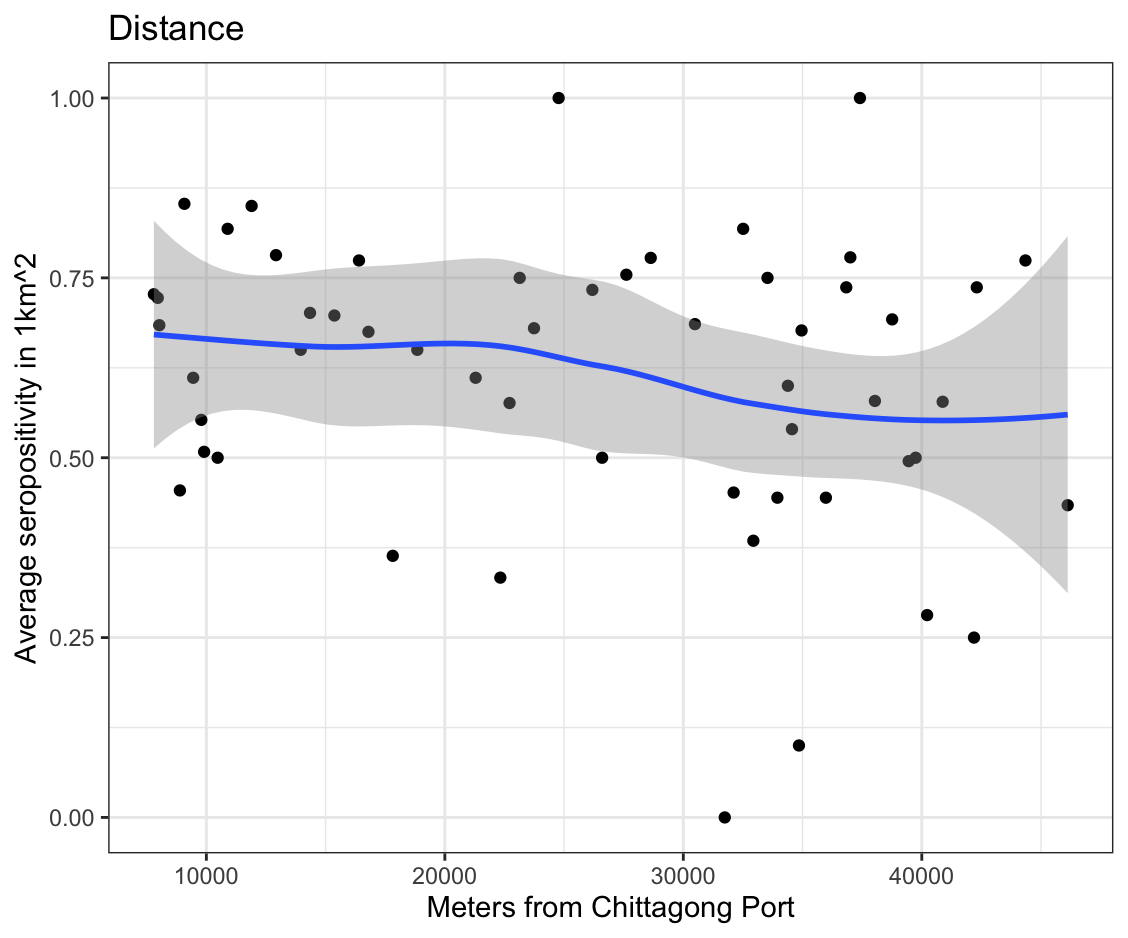
